# Mechanism of recurrent outbreak of the scarlet fever epidemics in mainland China

**DOI:** 10.1101/2024.08.09.24311775

**Authors:** Meirong Sun, Meijia Li, Naem Haihambo, Huiwen Zhang, Zhizhen Zhang, Xixi Zhao, Bin Wang, Mingrou Guo, Chuanliang Han

**Author notes:** **Corresponding author:** Chuanliang Han. Equally contributed (Meirong Sun, Meijia Li, Naem Haihambo).

## Abstract

In mainland China, most infectious diseases occur once a year, except for scarlet fever, which has been steadily breaking out twice a year in recent years. Over the years, the recurrence of scarlet fever, a contagious disease characterized by a distinctive red rash, has been a focus of attention. However, the oscillatory mechanism of scarlet fever remains unclear. This study aimed to uncover how meteorological factors contribute to the recrudescence of scarlet fever in mainland China. The study used a longitudinal public dataset covering 31 provinces in mainland China, containing 14 years of monthly scarlet fever infections, along with available local meteorological data. Power spectrum analysis was conducted on time series data, and correlation analyses were performed to assess the relationship between the oscillatory nature of epidemics and meteorological factors. We found that the scarlet fever epidemics generally occur twice a year in various provinces of China, and the timing of these outbreaks’ peaks progressively from southern to northern regions. Furthermore, we established an atlas that shows the relationship between scarlet fever oscillation and meteorological factors. Our findings indicated a significant correlation between the oscillation characteristics of scarlet fever in 50% of provinces and each natural meteorological factor. Our study presents a detailed description of the temporal and spatial changes in the oscillatory characteristics of scarlet fever for the first time and explores the oscillatory characteristics of natural meteorological conditions and their correlation with the number of scarlet fever infections. These findings could serve as a valuable guide for government prevention and control measures for the scarlet fever epidemic.

## Introduction

Scarlet fever, also known as scarlatina, is an acute respiratory infectious disease caused by group A hemolytic streptococcal bacteria [1]. The disease is spread through direct person-to-person transmission via airborne droplets, saliva, and wounds. Patients and carriers are the main sources of infection. Crowded places, such as schools and daycare centers can facilitate transmission. Scarlet fever can affect individuals of all age groups worldwide, although it is most prevalent among children aged 5 to 15 years. Children under the age of 3 are less likely to contract this condition [2–5].

An important feature of infectious diseases is their recurrent nature [6–8], which shows significant periodic oscillations over time [9–14]. In mainland China, most infectious diseases have annual outbreaks, with specific diseases having their own distinct outbreaks [9,10]. Scarlet fever, in particular, displays distinct oscillatory or recurring outbreak characteristics. Unlike other infectious diseases, such as influenza, which typically break out once a year, scarlet fever breaks out twice a year. The underlying mechanism driving its oscillatory infection remains unclear. The repeated outbreaks of scarlet fever have become a public health concern [15]. There have been some preliminary studies on the temporal dynamics of scarlet fever in various countries and regions around the world, including the United Kingdom [16–19], the Netherlands [20], South Korea [21], and Chinese Hong Kong[22–24]. In mainland China [25,26], previous studies have also delved into more precise geographical locations at a provincial-level [27–29], or city-level [4,28,30–33]. However, a systematic examination of scarlet fever’s oscillatory characteristics is still lacking.

The occurrence of scarlet fever’s epidemic form is determined by multiple factors, including environmental conditions, the nature and prevalence of the microorganism, distribution factors, and host population resistance. Among the environmental factors, climate and seasons are particularly significant. Notably, scarlet fever is observed in both tropical and temperate regions, and its incidence roughly correlates with the geographical location of a given country[30]. This correlation can be attributed to meteorological factors. Natural meteorological factors play an important role, affecting the transmission of infectious diseases[7,34,35]. Specifically, temperature (°C), precipitation (mm), humidity (%), and sunshine hours (h) are commonly studied meteorological factors that have been associated with many diseases, such as epidemic hemorrhagic fever [36–41], malaria [34,42–54], rabies [55], plague [56,57], and cholera [12,58]. These meteorological factors are frequently available in public data, allowing for the examination of their relationship with diseases. Some initial research has investigated the relationship between scarlet fever and meteorological factors in Chinese Mainland[4,28,59,60], even delving into possible connections between scarlet fever and pollution[61,62]. For example, a study focusing on the Beijing region of China revealed that the incidence of scarlet fever peaks between May and June (spring to early summer), accompanied by minor surges in incidence from November to early January (mid-autumn to mid-winter) [63]. Although these previous studies have explored the relationship between meteorological factors, environmental factors, and scarlet fever, the contribution of these natural factors to the oscillation of scarlet fever is currently unclear. China has notably diverse climate conditions, with a distinctive feature being the stark variation between regions. The northern part of the country has a “subarctic” climate, characterized by colder temperatures, while the southern regions are predominantly influenced by tropical weather patterns. This stark contrast makes China an excellent region for investigating the impact of meteorological factors on scarlet fever outbreaks.

If meteorological factors predominantly influence scarlet fever, it is reasonable to propose a hypothesis: the characteristics of infectious disease oscillations will exhibit significant shifts with changes in regional location. This shift is likely to be related to latitude or longitude, and the geographical location of a region on Earth is closely related to its corresponding climatic conditions, thereby affecting the number of infections of related infectious diseases, such as scarlet fever.

Therefore, in this study, we used public data from the China Public Health Science Data Center on scarlet fever in 31 provinces of China and the meteorological data of 31 provinces published in the China Statistical Yearbook spanning over 14 years to provide a detailed account of the temporal and spatial changes of the periodic characteristics of scarlet fever infections in mainland China, and to explore the oscillatory characteristics of natural meteorological conditions and their relationship with the number of people infected with scarlet fever. A research paradigm for analyzing oscillations will also be established to explore the relationship between the characteristics of scarlet fever oscillations and natural meteorological factors.

## Methods

### Data and Sources

Time series data on all available monthly reported and confirmed cases of scarlet fever in 31 administrative regions (23 provinces, 5 autonomous regions, 4 municipalities, and 2 special administrative regions, excluding Taiwan, Hong Kong, and Macau due to unavailable data) of mainland China from 2005 to 2018 were obtained from the Data Center of the China Public Health Science (Chinese Center for Disease Control and Prevention). We did not include recent years’ data due to the impact of COVID-19 and the resulting lockdowns in China, which have significantly affected the natural property of the temporal dynamics of scarlet fever.

Monthly reported data of meteorological elements, including temperature, precipitation, humidity and sunshine hours, were obtained for 31 provinces in mainland China from January 2005 to December 2018 were obtained from the China Statistical Yearbook 2006–2019. The meteorological data included in this study are continuous values that vary with time, which directly reflect the actual natural conditions each month. This dataset is publicly available worldwide and is reported annually.

### Ethical Considerations

This study used public data from the China Public Health Science Data Center and the China Statistical Yearbook. Our study did not involve any interventions with human participants. This study was approved by the Ethics Committee of Beijing Sport University (2022142H), China.

### Power Spectrum Analysis

Data processing was performed using custom scripts in MATLAB (www.mathworks.com). Spectrum analysis was utilized to quantify fluctuations and the recurrence of epidemics for scarlet fever (Fig 1C) and the natural factors (Fig 2). Similar methods have been applied in various biomedical fields, such as life sciences [64–66], neuropsychological disorders [67–69], infectious diseases [11,70–73], etc. The power spectral density (PSD) for each infectious disease was computed using the multi-taper method with the Chronux toolbox[74], an open-source, data analysis toolbox (Chronux) available at http://chronux.org.

**Figure 1.**
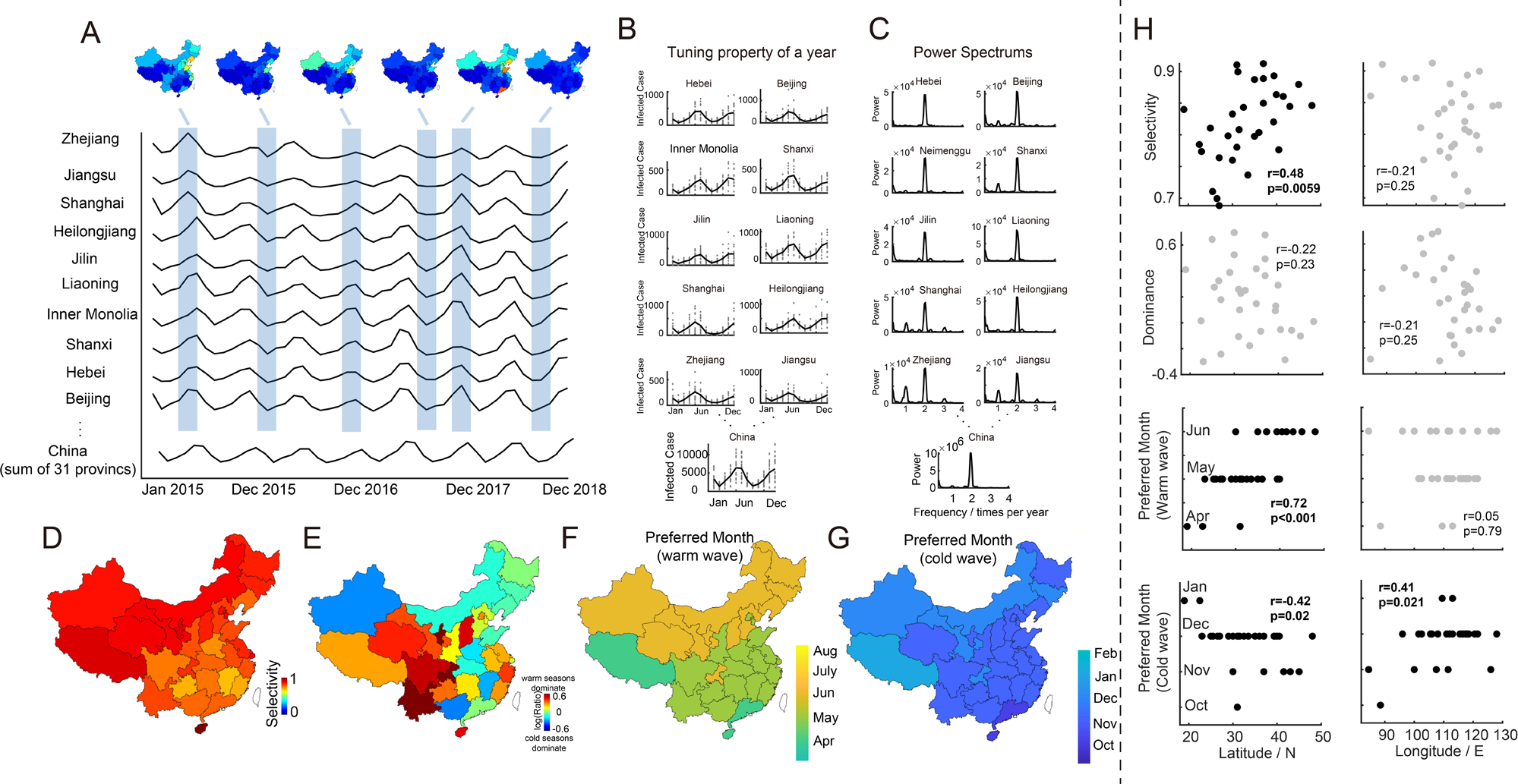
Periodic phenomena of scarlet fever epidemics with tuning curve, power spectrum and topographic map of several indices. A. Monthly incidence of scarlet fever infections in 10 representative provinces, accompanied with topographic maps depicting infected case distribution at specific intervals. B. Tuning curves of scarlet fever epidemics in example provinces. C. Spectrogram of scarlet fever epidemics from January 2005 to December 2018 in the aforementioned example provinces. D. Geospatial distribution of the selectivity of scarlet fever epidemics across provinces. E. Geospatial distribution of the dominance level of scarlet fever epidemics for each province. F. Geospatial distribution of the peaking month of scarlet fever epidemics in warm seasons for each province. G. Geospatial distribution of the peaking month of scarlet fever epidemics in cold seasons for each province.

**Figure 2.**
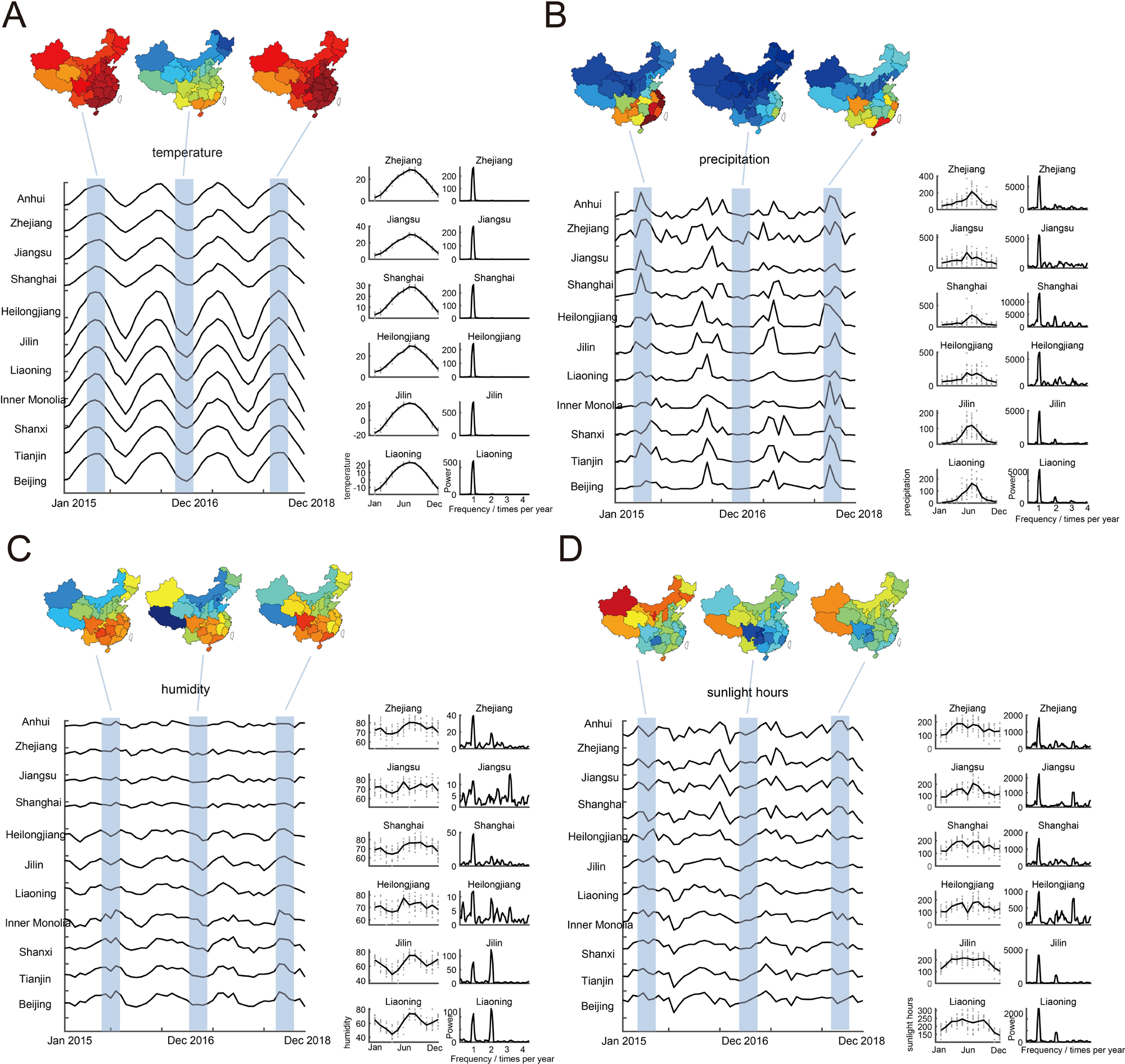
Oscillatory patterns (power spectrum and tuning curves) of various meteorological elements. Panels A–D show the monthly time series of meteorological elements (temperature, precipitation, humidity, and sunshine hours) and their spectrogram from January 2005 to December 2018 in China. The geospatial distribution of the infected cases in 31 provinces was shown in the topographic map above each panel. The tuning curves and power spectrum of time series data are shown on the right side in example provinces.

### Tuning curves

The tuning curve of the monthly infected cases illustrates an essential profile of the outbreak of scarlet fever in mainland China (Fig 1B), which gives a direct monthly snapshot of the situation based on historical data. We assumed that all infectious diseases included in this study follow a similar annual trend each year considered in this study, as previously researched [9,11]. Thus, we took the monthly average number of infected cases and computed them into a tuning curve (Eq 1). Each infectious disease in this study has a tuning curve, revealing a clear oscillatory pattern within a year. This method is also employed to measure the tuning curve of meteorological elements (Fig 2).

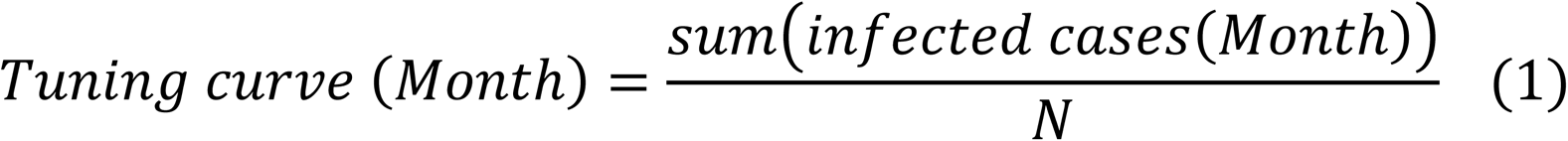

where N is the number of the year.

### Preferred month and selectivity of the epidemic outbreak

Two disease indices were defined: preferred month and infection selectivity (Fig 1D-G), which are important indicators that show the infectious property of the epidemics caused by the disease in a year. The preferred month index represents the month with the highest number of infection cases in a year. The selectivity index for infection is defined as 1 minus the ratio of the minimum and the maximum number of infected cases in a year. A higher selectivity index (closer to 1) indicates that outbreaks are concentrated within specific months, while a lower index (closer to 0) implies a more year-round occurrence.

### Data Filtering

To preserve the most obvious periodic information, the original continuous data were high-pass filtered 2.5 times per year and low-pass at 1.5 times per year (Fig 4A). Both the high-pass and low-pass filters were zero-phased FIR filters (third-order Butterworth filter), which filter the data in both forward and backward directions to nullify any phase delays introduced by each filter.

### Correlation Analysis

The Pearson correlation was used to measure the relationship between properties of the tuning curve and the locations of each province (Fig 1H). Additionally, Pearson correlations were used to evaluate the relationship between infected cases (Fig 3; also for filtered data in Fig 4) and meteorological elements, respectively.

**Figure 3.**
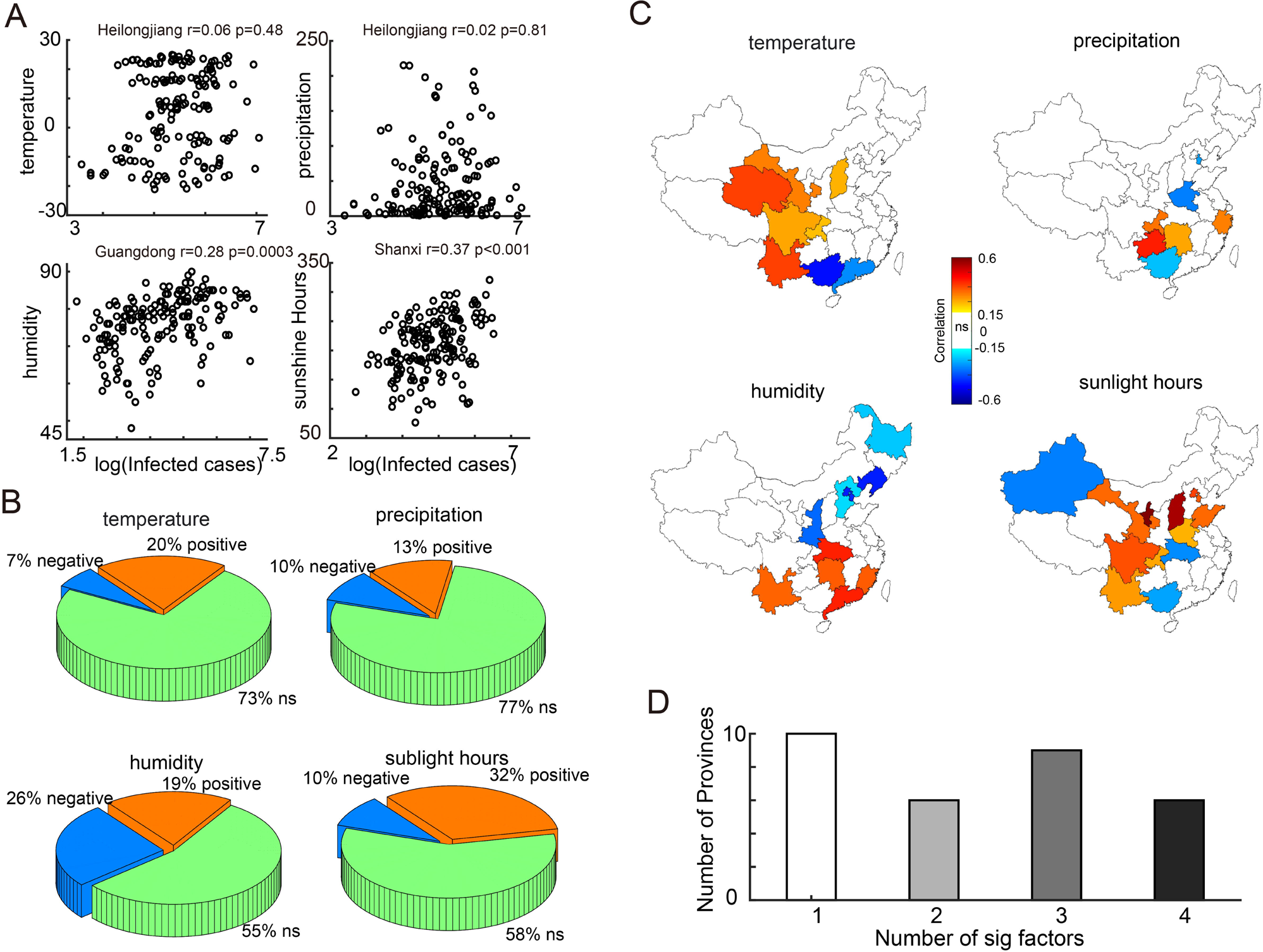
Relationship between infected cases of scarlet fever epidemic and meteorological elements. A. Scatter plots illustrating the number of cases of scarlet fever in some example provinces and temperature, precipitation, humidity, and sunshine hours, respectively (Heilongjiang: northeast region, Guangdong: south region, Shanxi, central region). B. Pie chart shows the proportion of different types of correlations between scarlet fever cases and four meteorological elements (Orange for positive correlation, blue for negative correlation, green for no significant correlation). C. Geospatial distribution of scarlet fever epidemics in provinces with significant correlations to specific meteorological elements. D. Count of provinces related to different significant numbers (from 1 to 4) of meteorological factors.

**Figure 4.**
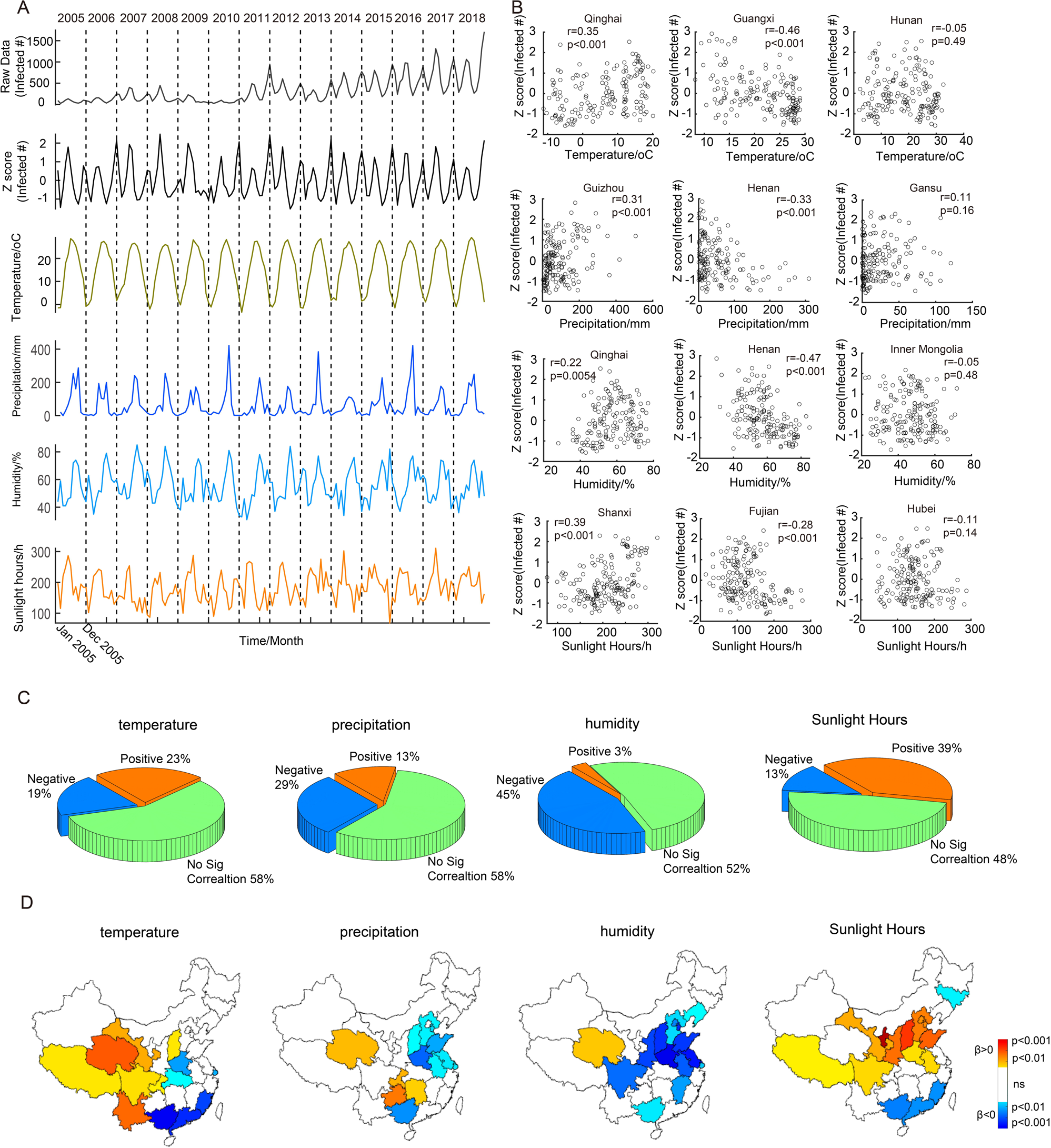
Relationship between oscillation characteristics of scarlet fever epidemic and meteorological elements. A. The monthly incidences of scarlet fever, associated filtered time series data and four meteorological elements (shown in different colors) in Shandong (as an illustrative example). B. Scatter plots illustrate the number of cases of scarlet fever in some example provinces and temperature, precipitation, humidity, and sunshine hours, respectively (Central region: Shanxi, Henan, Hubei; southwest region: Qinghai, Guizhou; North region: inner Mongolia, Gansu; Southeast region: Fujian). C. Pie chart shows the proportion of different types of correlations between scarlet fever oscillations and four meteorological elements (Orange for positive correlation, blue for negative correlation, and green for no significant correlation). D. Geospatial distribution of scarlet fever oscillations in provinces that have significant correlations with specific meteorological elements.

### Statistical Methods

Multiple t-tests with Bonferroni correction were used to compare the oscillation characteristics before and after 2011 (Fig 5).

**Figure 5.**
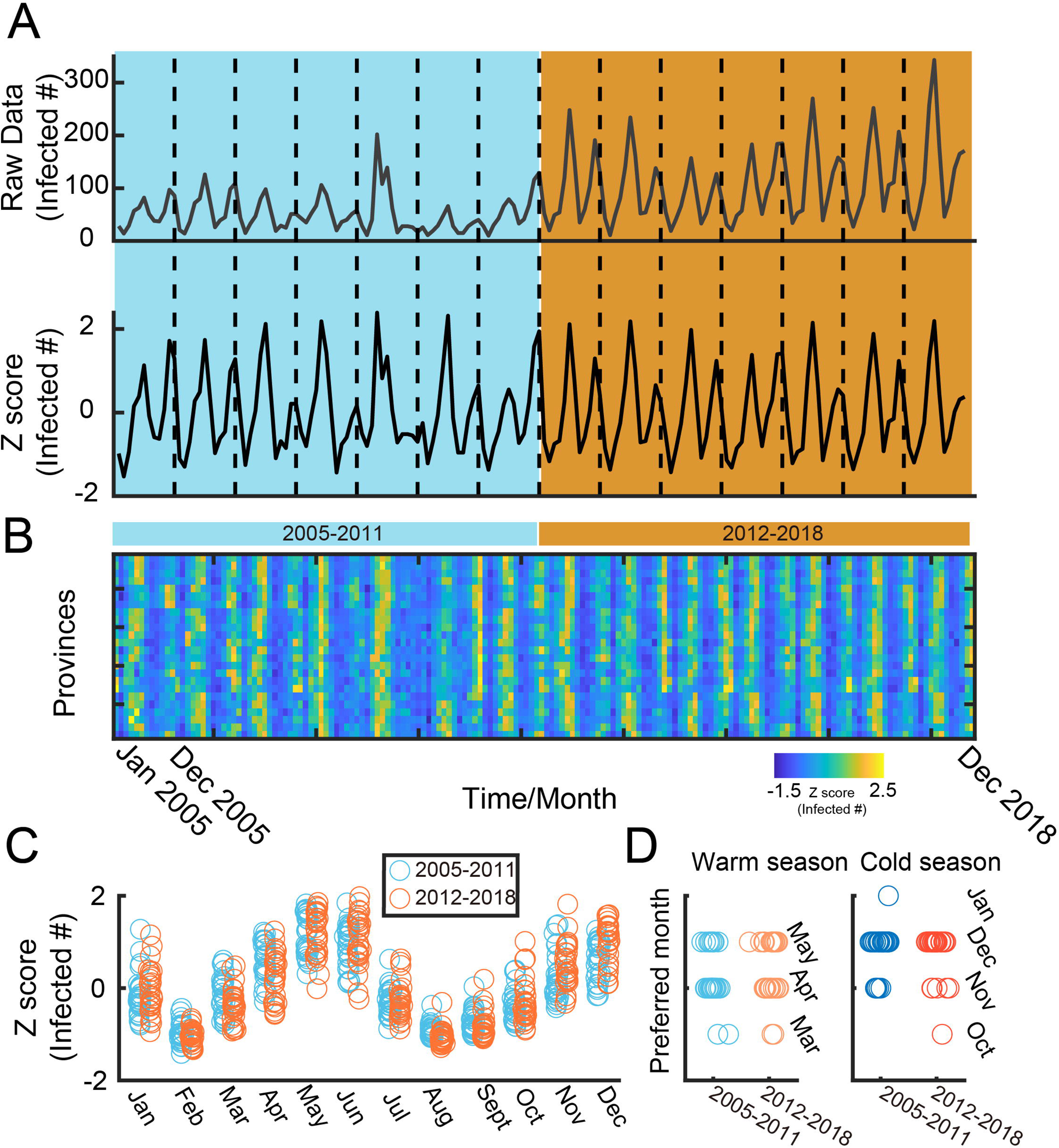
Oscillation characteristics before and after 2011. A. The monthly incidences of scarlet fever, associated filtered time series data in Gansu (as an illustrative example). B. The monthly incidences of scarlet fever, associated filtered time series data in multiple provinces (n=25). C. The blue dots indicate the number of infected cases every month in year from 2005-2011, while orange dots indicate that in year from 2012-2018. D. The peaking month of scarlet fever epidemics in warm and cold seasons for each province in year from 2005-2011, while orange dots indicate that in year from 2012-2018.

## Results

### Spaciotemporal patterns of scarlet fever incidences in mainland China

This study examined monthly data of confirmed cases of scarlet fever in31 provinces in China spanning from January 2005 to December 2018 (Fig 1). Figure 1A shows the time series data of scarlet fever in 10 representative provinces, with the topographic map displayed above. Evidently, each province displays distinct oscillatory patterns in its scarlet fever outbreaks as depicted by their tuning curves (Fig 1B) and power spectrum (Fig 1C). These oscillations manifest with different peaks, occurring once, twice, or even three times a year. Notably, the power spectrum prominently highlights the occurrence of a biannual peak as the most significant pattern.

Through an in-depth analysis of oscillation characteristics, we observed a decreasing trend in the selectivity of scarlet fever from northwest to southeast (Fig 1D). Scarlet fever has biannual outbreaks, with peaks occurring during both the warm (April to September) and cold seasons (October to March). Then, the dominance level was calculated as the ratio of the number of cases in the warm peak and cold peak. The results indicate that in the southwest region, more infections occur during the warm season, while in the northeast region, more infections occur during the cold season (Fig 1E). Further, we compute the preferred month during both the warm and cold seasons (Fig 1FG). Intriguingly, our analysis revealed that the timing of scarlet fever’s peak in the southeast consistently precedes that in the northwest region. Furthermore, to verify the statistical significance of the shift, we conducted Pearson correlations between the properties (Fig 1D-F) and the provinces’ locations (latitudes and longitudes, Fig 1H). The selectivity is positively correlated with the latitude but not longitude, while the dominance level does not correlate with the locations. The preferred month in warm seasons is positively correlated with the latitude but not with the longitude, but the preferred month in cold seasons is positively correlated with both latitude and longitude.

### Periodic phenomena of meteorological elements in 31 provinces in China

Through the display of the oscillation characteristics, especially the temporal characteristics of scarlet fever, our findings reveal a gradual change from south to north or vice versa among various provincial capitals in China. This indicates that natural meteorological factors can potentially influence the incidence of scarlet fever, due to the substantial climate variations between northern and southern China. Therefore, we further explored the oscillatory characteristics of various climate factors (Fig 2) and then conducted a power spectrum analysis of the time series data encompassing temperature, precipitation, humidity, and sunshine hours across 31 provinces in China (Fig 2A-D). Notably, all four meteorological elements in most provinces in China exhibit evident oscillatory patterns over time. Temperature and precipitation follow a yearly cyclical pattern, while humidity and sunshine hours may exhibit either strong or weak biannual periodicity. These outcomes suggest a potential correlation between epidemics in certain provinces and these meteorological elements.

### Relationship between the scarlet fever infection and meteorological elements in China

From the observation of the average infected cases of the scarlet fever epidemic and the average meteorological elements, scarlet fever infections may be influenced by several natural factors. Therefore, we then investigated the relationship between the scarlet fever outbreaks and meteorological elements in each province in China based on correlation analyses.

Several scatter plots are shown in Figure 3A. For different provinces, we found that there are different associations between the number of people infected with scarlet fever and natural factors, indicating that distinct regions may require tailored strategies to address the same disease. This diversity is further illustrated in Figure 3B. In China, 27% of provinces exhibit a correlation between scarlet fever outbreaks and temperature (with 20% positive correlation and 7% negative correlation), while 23% of provinces exhibit a correlation between scarlet fever outbreaks and precipitation (with 13% positive correlation and 10% negative correlation). Furthermore, 45% of provinces show a correlation between scarlet fever outbreaks and humidity (with 19% positive correlation and 26% negative correlation), and 42% of provinces display a correlation between scarlet fever outbreaks and sunshine hours (with 32% positive correlation and 10% negative correlation).

Specifically, the topographic map (Fig 3C) highlights the regional distribution. Notably, provinces with a significant positive correlation between scarlet fever epidemics and *t*emperature are clustered in the southwest region, while those with a significant negative correlation with temperature are found in the southern region. The influence of precipitation appears to be relatively minimal, showing the least impact, as indicated by the provinces involved. Positive correlations with precipitation are mainly found in the central and southern regions, while sporadic provinces with negative correlation in the southern and northern regions. Provinces that are positively correlated with humidity are mainly situated in the southern regions, while those that are negatively correlated with humidity are mainly in north and northeast areas of China. Furthermore, the provinces displaying a positive correlation with daylight hours are primarily found in the southwest and north regions, while those with a negative correlation are mainly found in the northwest and central southern regions.

In sum, scarlet fever outbreaks are affected by various meteorological types across most provinces in China, and this impact cannot be attributed to a single factor, but rather a combination of factors (Fig 3D). Further analysis revealed that only 10 provinces were affected by a single meteorological factor, while more provinces were affected by multiple factors concurrently.

### Relationship between the periodic characteristics of scarlet fever infection and meteorological elements in mainland China

In the analysis above, we found that scarlet fever epidemics are closely related to natural meteorological factors. However, simply analyzing the correlation between the number of infected cases and meteorological indicators does not fully explain the role of periodic oscillations in this context. To tackle this problem, we first filter the original continuous time series data of scarlet fever infections, retaining the data with a biannual frequency. Next, we standardize the filtered data on a yearly basis by calculating the z-score, considering that the amplitude of outbreaks varies from year to year. This process ensured comparability even when dealing with data from different scales. Then trend information was removed, retaining only the periodic information of the original data retained (Fig. 4A first two rows). We then analyzed the correlation between the *z*-score value and various meteorological indicators. Similarly, the results indicate diverse correlations between the *z*-score and meteorological factors across different provinces (Fig 4B).

When considering the entire country, more provinces display significant outcomes, especially the influence of temperature, which is more pronounced (Fig 4C). Specifically, 42% of provinces exhibit a correlation between scarlet fever oscillations and temperature (with 23% showing positive correlation and 19% showing negative correlation), while another 42% of provinces are observed to show a correlation between scarlet fever oscillations and precipitation (with 13% positive correlation and 29% negative correlation). Additionally, 48% of provinces reveal a correlation between oscillations and humidity (with 3% positive correlation and 45% negative correlation), and 52% of provinces display a correlation between scarlet fever oscillations and sunshine hours (with 39% positive correlation and 13% negative correlation).

Specifically, regarding regional distribution (Fig 4D), provinces in the southwest region show significant positive correlations between scarlet fever oscillations and temperature, while those in the central and southern regions exhibit significant negative correlations with temperature. The provinces that are positively correlated with precipitation are mainly located in the central and southern regions, while the northern region mainly shows a negative correlation to the precipitation. Provinces that are positively correlated with humidity are mainly observed in Qinghai in the western region, while those that are significantly negatively correlated with temperature are concentrated in northern China. Finally, provinces that are positively correlated with sunshine hours are mainly located in the central region, while those that are significantly negatively correlated with temperature are concentrated in the northeastern and southern regions. In sum, after removing the trend information and retaining only the periodic information from the original data, some significant correlations became insignificant while some others became insignificant. However, this result does not conflict with the results of Figure 3 or flip polarity.

### Periodic characteristics of scarlet fever infection remains consistent before and after 2011

In 2011, the incidence of scarlet fever in China experienced a significant increase, marking a turning point. This phenomenon has been reported in previous studies and is thought to be related to changes in the natural characteristics of the virus (ref). Our results also demonstrate obvious phenomena (Fig 4A, Fig 5A, B), but the changes in oscillation characteristics have not been studied. Using the normalized method shown in Figure 4A, it is clear that the oscillatory phenomenon is comparable before and after 2011. This is also evident in the tuning curves (Fig 5C). The two peaking months of epidemic in warm and cold seasons respectively also did not show a significant difference (Fig 5D). This indicates that although the number of people infected with scarlet fever increased in 2011, its natural oscillation characteristics remained unchanged.

## Discussion

### Principal Findings

This study presents the first endeavor to meticulously examine the temporal and spatial changes in the oscillation of scarlet fever outbreaks. We explored the oscillatory characteristics of natural meteorological conditions and their correlation with scarlet fever infections. Additionally, we established a research framework for analyzing oscillations to explore the relationship between the oscillatory characteristics of scarlet fever and natural meteorological factors. Our study not only advances our understanding of these oscillatory patterns but also holds practical implications for effective public health management and prevention measures, providing a valuable resource for local authorities.

First and foremost, the temporal patterns observed present a compelling spatial progression of scarlet fever incidences. Specifically, varied time lags are observed across different provinces, with the peak of scarlet fever starting from the south and then spreading northward (Fig 1). This phenomenon has the potential to serve as a predictive indicator, assisting northern provinces in preemptively addressing and managing the emerging epidemic. We found a significant shift in the oscillation characteristics of scarlet fever from north to south in China (Fig 1H), which validates the hypothesis we proposed in the introduction. This also confirms that meteorological factors have a tremendous impact on the epidemic of scarlet fever in China. This research sample can be expanded to other countries worldwide.

Moreover, our study shows a correlation between local scarlet fever oscillation characteristics and meteorological factors in different provinces of China (Fig 3 & 4). Notably, factors such as humidity (45%) and sunshine hours (42%) have a higher chance of correlation with scarlet fever outbreaks, which that aligns with previous research. After removing the trend information and retaining only the periodic information from the original data, more provinces display significant outcomes for all four factors, especially the influence of temperature, which is more salient (27% to 42%).

Above all, our research reveals the heterogeneous nature of the relationship between different provinces and meteorological influences. This revelation highlights the imperative for a nuanced and individualized approach to epidemic prevention. Instead of adopting a uniform strategy across all regions, our study emphasizes the critical importance of discerning and accounting for the unique factors that characterize each region. By doing so, authorities can ensure that their preventive measures are not only effective but also tailored to the specific exigencies of the region in question. In sum, our study extends on current knowledge regarding scarlet fever dynamics by unraveling the intricate relationship between temporal and spatial factors and meteorological conditions. By aligning public health measures with specific meteorological conditions, local governments can more effectively customize their interventions to each unique context, or during the colder months when schools resume.

### Comparison With Prior Work

To our knowledge, our study is the first to illustrate the spatiotemporal dynamics of scarlet fever from an oscillatory view. Previous studies have provided valuable insights into the characteristics and spatiotemporal distribution of scarlet fever and have even included meteorological variables [75]. These studies have, however, primarily focused on fundamental descriptive analysis [25] rather than providing a detailed characterization and explanation of periodic features. Some earlier studies attempted to directly characterize the oscillation of scarlet fever [9,11], but these often utilized national-level data, lacking the granularity of provincial refinement. For example, Mahara et al., (2016), found significant correlations between temperature and relative humidity in Beijing [30].

Furthermore, it is worth noting that the precision of the data employed primarily encompasses annual occurrences[61], largely overlooking the potential influence stemming from meteorological factors that can vary significantly within a single year. Thus, there was little discussion regarding the contribution of natural meteorological factors on the oscillation characteristics of scarlet fever over multiple geographical areas. Previous research has generally focused on infectious diseases in a vaguer and less targeted way in terms of geography and not on oscillations. Our findings underscore the importance of considering meteorological factors in the context of disease oscillations, shedding light on potential links between climate variables and scarlet fever dynamics. The study presents a data analysis paradigm for studying the oscillation of infectious diseases (Fig 4). This can be used by future researchers to further explore various in-depth mechanisms of infectious disease oscillations.

### Limitations

Our study has some limitations that should be addressed in future research. Firstly, the availability of more detailed spatial-scale data is currently a challenge. While our analysis covered all 31 provinces in China, the lack of finer-scale data prevents us from conducting a more in-depth investigation of scarlet fever outbreaks within individual cities. This limitation underscores the need for improved data collection and sharing mechanisms. Additionally, the absence of consideration for population migration between provinces within China is another limitation. Despite the scarlet fever population is mainly in children, we assume that their migration is much weaker than that of adults. However, future research should still take this factor into account.

### Conclusion

In conclusion, our study represents a significant advancement in the understanding of scarlet fever dynamics, particularly in terms of its spatiotemporal oscillations. We have highlighted the importance of considering meteorological factors in disease oscillation studies and introduced a comprehensive data analysis paradigm that can be applied to investigate similar phenomena in other infectious diseases. This work provides a foundation for future investigations to delve deeper into the intricate mechanisms steering disease oscillations. It can ultimately contribute to more effective public health interventions.

## Data Availability

This study used public data from the China Public Health Science Data Center and the China Statistical Yearbook.

## Acknowledgments

This work was funded by the Fundamental Research Funds for Central Universities (2021TD010), Open project of Beijing Key Laboratory of Mental Disorders (2020JSJB02), the National Natural Science Foundation of China (82201701), Beijing Municipal Hospital Research and Development Project (PX2021068), Advanced Innovation Center for Human Brain Protection Project (3500-12020137)

## Contributors

CH, ML, XZ conceived and designed the study. CH, ML, NH, BW, MS contributed to the literature search, CH, ML contributed to data collection. CH contributed to the data analysis, and the interpretation of results. All authors contributed to writing the paper.

## Appendix-tables

**Table 1.**
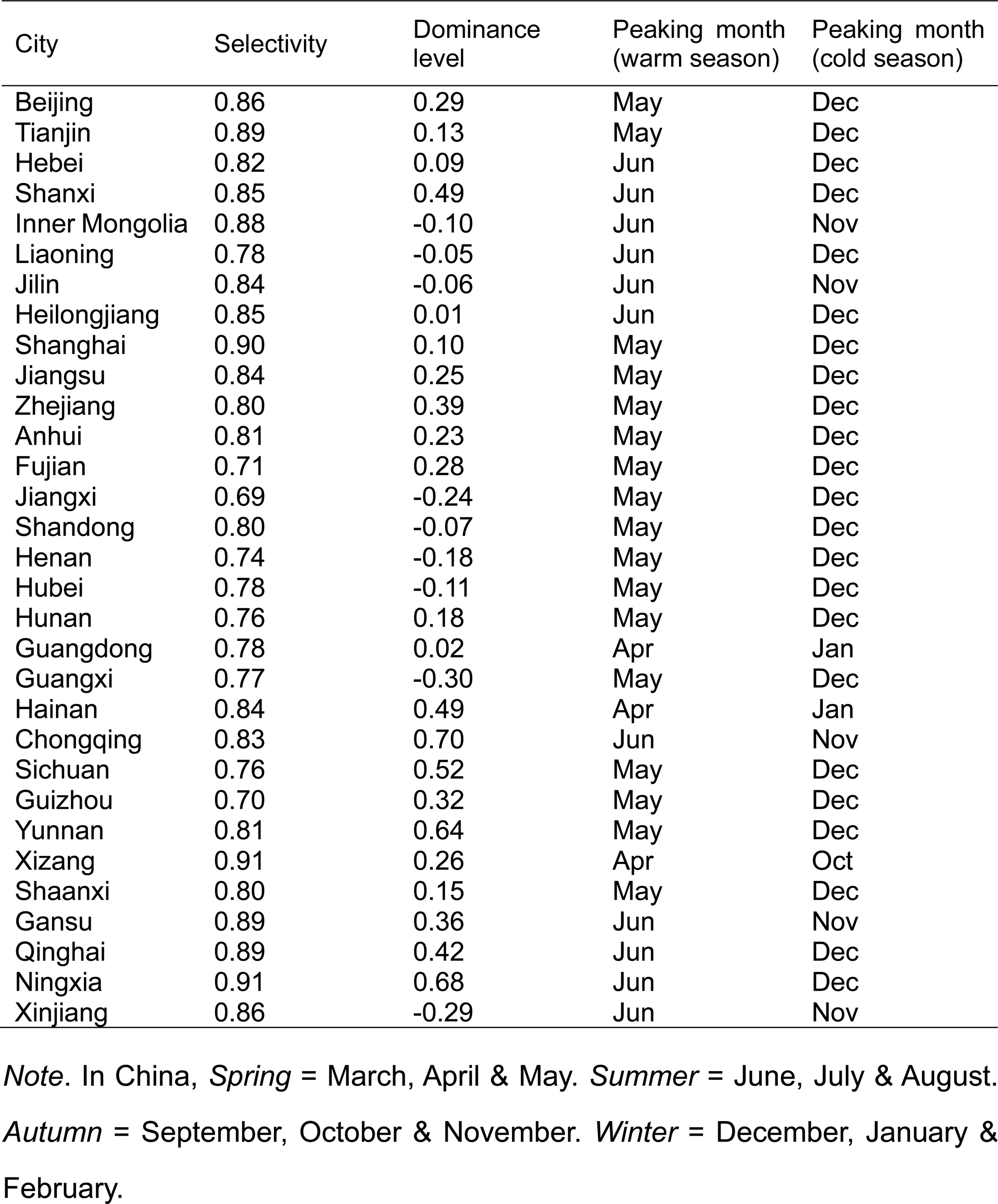
Descriptive statistics of periodic phenomena of scarlet fever epidemics.

**Table 2.** Relationship between infected cases of scarlet fever epidemic and meteorological elements.

|  | Tempearture |  | Percipitation |  | Humidity |  | Sunshine hours |  |
| --- | --- | --- | --- | --- | --- | --- | --- | --- |
|  | *r* | *p* | *r* | *p* | *r* | *p* | *r* | *p* |
| Beijing | 0.05 | 0.51 | 0.08 | 0.32 | 0.24 | 0.00 | 0.25 | 0.00 |
| Tianjin | -0.02 | 0.77 | 0.17 | 0.04 | 0.32 | 0.00 | 0.23 | 0.00 |
| Hebei | 0.09 | 0.25 | 0.11 | 0.16 | 0.12 | 0.11 | 0.11 | 0.17 |
| Shanxi | 0.16 | 0.04 | 0.12 | 0.14 | 0.24 | 0.00 | 0.37 | 0.00 |
| Inner Mongolia | 0.01 | 0.85 | 0.05 | 0.54 | 0.02 | 0.75 | -0.05 | 0.52 |
| Liaoning | -0.04 | 0.61 | 0.13 | 0.08 | 0.20 | 0.01 | -0.02 | 0.77 |
| Jilin | 0.01 | 0.89 | 0.09 | 0.23 | 0.12 | 0.11 | -0.03 | 0.66 |
| Heilongjiang | 0.06 | 0.48 | 0.02 | 0.81 | 0.07 | 0.39 | -0.08 | 0.32 |
| Shanghai | -0.15 | 0.05 | 0.00 | 0.98 | 0.06 | 0.47 | -0.06 | 0.43 |
| Jiangsu | -0.07 | 0.36 | 0.02 | 0.82 | 0.12 | 0.11 | 0.04 | 0.60 |
| Zhejiang | -0.03 | 0.69 | 0.20 | 0.01 | 0.08 | 0.28 | -0.10 | 0.18 |
| Anhui | -0.06 | 0.48 | 0.00 | 1.00 | 0.05 | 0.52 | -0.04 | 0.63 |
| Fujian | -0.10 | 0.21 | 0.02 | 0.83 | 0.21 | 0.01 | -0.13 | 0.10 |
| Jiangxi | -0.06 | 0.41 | 0.01 | 0.89 | 0.04 | 0.57 | -0.04 | 0.63 |
| Shandong | -0.08 | 0.30 | 0.15 | 0.06 | 0.17 | 0.02 | 0.22 | 0.00 |
| Henan | -0.12 | 0.12 | 0.20 | 0.01 | 0.27 | 0.00 | 0.16 | 0.04 |
| Hubei | -0.11 | 0.14 | 0.03 | 0.66 | 0.30 | 0.00 | -0.19 | 0.02 |
| Hunan | -0.03 | 0.74 | 0.17 | 0.03 | 0.27 | 0.00 | -0.09 | 0.23 |
| Guangdong | -0.19 | 0.01 | 0.02 | 0.84 | 0.28 | 0.00 | -0.09 | 0.27 |
| Guangxi | -0.29 | 0.00 | 0.15 | 0.05 | 0.02 | 0.80 | -0.17 | 0.03 |
| Hainan | -0.11 | 0.52 | 0.09 | 0.62 | 0.08 | 0.65 | 0.03 | 0.87 |
| Chongqing | 0.15 | 0.05 | 0.21 | 0.01 | 0.09 | 0.24 | 0.17 | 0.03 |
| Sichuan | 0.17 | 0.03 | 0.02 | 0.79 | 0.19 | 0.01 | 0.24 | 0.00 |
| Guizhou | 0.11 | 0.14 | 0.28 | 0.00 | 0.23 | 0.00 | -0.07 | 0.38 |
| Yunnan | 0.25 | 0.00 | 0.15 | 0.07 | 0.00 | 0.95 | 0.18 | 0.02 |
| Xizang | 0.13 | 0.10 | 0.00 | 0.99 | 0.12 | 0.14 | 0.08 | 0.29 |
| Shaanxi | 0.04 | 0.58 | 0.09 | 0.27 | 0.24 | 0.00 | 0.14 | 0.08 |
| Gansu | 0.20 | 0.01 | 0.11 | 0.16 | 0.06 | 0.44 | 0.22 | 0.00 |
| Qinghai | 0.25 | 0.00 | 0.12 | 0.14 | 0.16 | 0.04 | 0.06 | 0.45 |
| Ningxia | 0.33 | 0.12 | 0.02 | 0.92 | 0.24 | 0.25 | 0.55 | 0.01 |
| Xinjiang | -0.06 | 0.47 | 0.16 | 0.06 | 0.15 | 0.07 | -0.20 | 0.02 |

*Note.* *r* = correlation coefficient (Pearson), *p* = significance level.

**Table 3.**
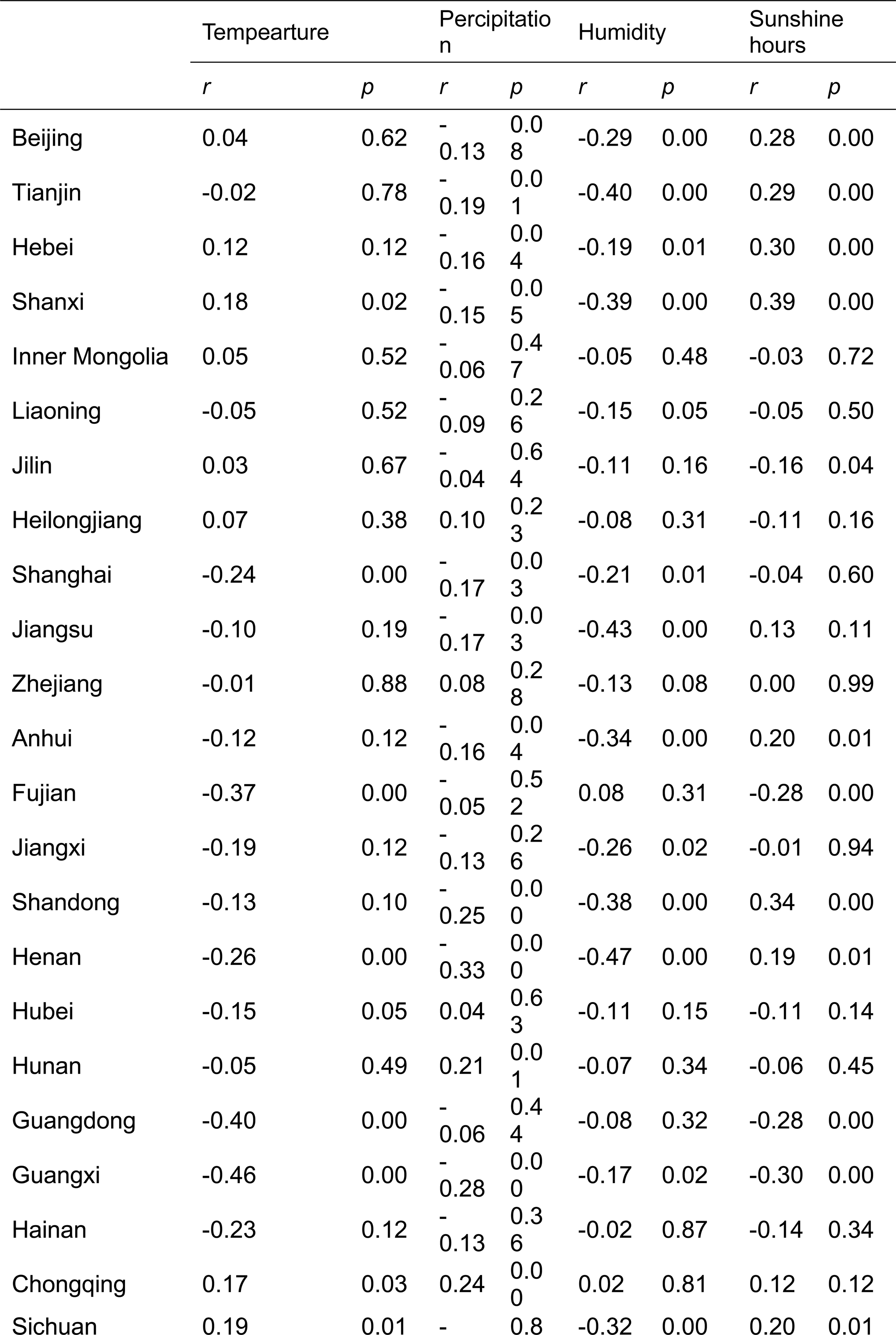
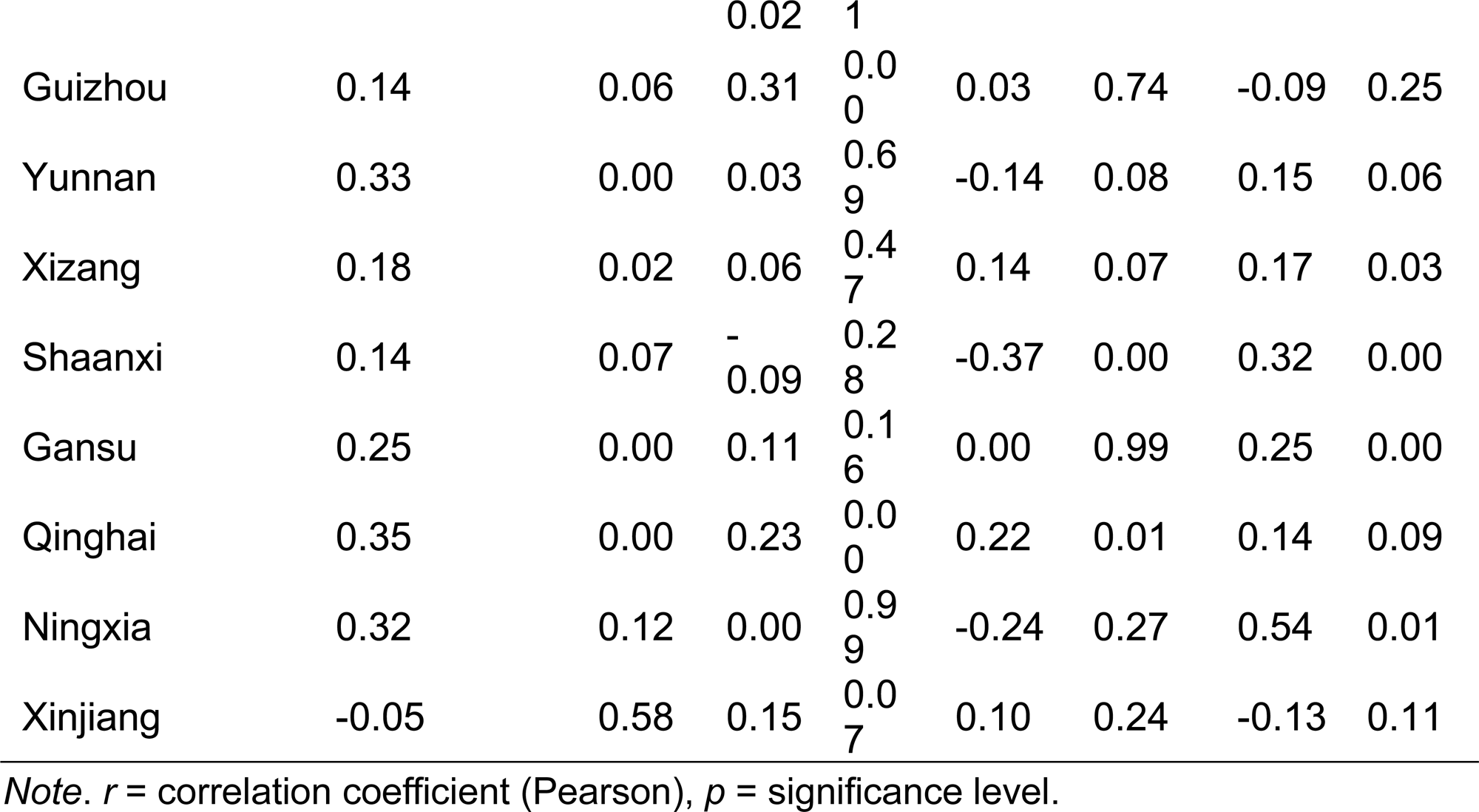
Relationship between oscillation characteristics of scarlet fever epidemic and meteorological elements.

